# Development and validation of a modified Cambridge Multimorbidity Score for use with internationally recognized electronic health record clinical terms (SNOMED CT)

**DOI:** 10.1101/2022.03.02.22271765

**Authors:** Ruby S. M. Tsang, Mark Joy, Heather Whitaker, James P. Sheppard, John Williams, Julian Sherlock, Nikhil Mayor, Bernardo Meza-Torres, Elizabeth Button, Alice J. Williams, Debasish Kar, Gayathri Delanerolle, Richard McManus, F. D. Richard Hobbs, Simon de Lusignan

## Abstract

**Background:** People with multiple health conditions are more likely to have poorer health outcomes and greater care and service needs; a reliable measure of multimorbidity would inform management strategies and resource allocation. This study aims to develop and validate a modified version of the Cambridge Multimorbidity Score in an extended age range, using clinical terms which are routinely used in electronic health records across the world (SNOMED CT).

**Methods and Findings:** We curated new variables describing 37 health conditions and modelled the associations between these and 1-year mortality risk using the Cox proportional hazard model in a development dataset (n=300,000). We then developed two simplified models – a 20-condition model as per the original Cambridge Multimorbidity Score, and a variable reduction model using backward elimination with Akaike information criterion as the stopping criterion. The results were compared and validated for 1-year mortality in a synchronous validation dataset (n=150,000), and for 1-year and 5-year mortality in an asynchronous validation dataset (n=150,000).

Our final variable reduction model retained 21 conditions, and the conditions mostly overlapped with those in the 20-condition model. The model performed similarly to the 37- and 20-condition models, showing high discrimination and good calibration following recalibration.

**Conclusions:** This modified version of the Cambridge Multimorbidity Score allows reliable estimation using clinical terms which can be applied internationally across multiple healthcare settings.

## Introduction

Many epidemiological analyses, including measuring the impact of disease or the effectiveness of therapies, require a single measure of comorbidity. People with multiple health conditions are likely to have poorer health outcomes and require more intensive treatment and monitoring, placing significant and increasing demand across the spectrum of health services [1]. Evaluating multimorbidity is important in allocating resources, optimising management strategies, and facilitating research. This can be achieved through composite scores that quantify the effect of specific comorbid conditions on health service utilisation, unplanned hospital admission, and mortality [2, 3].

There have been a number of approaches to measuring comorbidity. The Charlson Comorbidity Index (CCI) is a commonly used composite morbidity score with condition weightings based on mortality [2]. However, the management of multimorbidity has seen a paradigm shift towards a greater focus on primary care and non-hospital management of disease [4–7]; the CCI, having been designed for use in secondary care and is based on secondary care coding systems, is not ideal for use in primary care. Moreover, the contribution of its twelve selected comorbidities since its validation in 1987 has changed, requiring the index to be re-evaluated and re-validated. Other approaches have included the number of comorbidities, though the weakness of this is the lack of weighting or to count the number of disease areas or risk groups.

To improve on these limitations, the Cambridge Multimorbidity Score (CMMS) was developed in 2020 for use in primary care practices, using data from the Clinical Practice Research Datalink (CPRD) [8]. The CMMS used 37 conditions (and 20 in its simplified form) to predict primary care consultations, unplanned hospital admissions, and death as primary outcomes. The weighting-based outcome-specific scores of the CMMS is reported to outperform the CCI across all three primary outcomes. However, the original analysis excluded patients under 21 years, which may limit its validity and utility in studies that include individuals outside of this age range.

The CMMS was originally developed and validated using comorbidities defined with Read clinical terminology, a thesaurus of clinical terms used to record patient findings and procedures in computerised medical records (CMR) [9]. Since April 2016 the Read terminology has not been updated. It was then retired from clinical use in English General Practice in 2018 and was replaced by the systematised nomenclature for medicine (SNOMED) clinical terms CT [10] which is used in electronic health records across the world. Potential benefits of SNOMED CT include its comprehensive nature, its capability to be machine processed, its precise collection of clinical terminology as well as its international implementation.

We conducted this study to develop and validate a modified version of the CMMS with an extended age range, which is solely based on SNOMED CT, and using routinely collected primary care data from the Oxford-Royal College of General Practitioners (RCGP) Research and Surveillance Centre (RSC).

## Methods

### Data source and variables

We use pseudonymised CMR data from the RSC sentinel network database, which is recruited to be representative of the general population. The UK has registration-based primary care in which each patient registers with a single general practice.

We included all patients who were registered for at least 12 months before the study start date, and aged 16 years and older at the study index date for each model. We split the cohort into three separate datasets (development set, validation set 1 with synchronous outcome, validation set 2 with asynchronous outcomes) (**Figure 1**) using block randomisation in the ratio of 2:1:1. To minimise the effect of random variation between practices on mortality, the cohort was separated into four subsets using the best linear unbiased estimator from a mixed effects logistic regression with age (standardised) and sex fixed effects and a practice random effect, prior to block randomisation (**S1 Figure**). We further applied similar inclusion/exclusion criteria for selecting individuals to those described in the original analysis [8] (**S2 Figure**). We then randomly sampled 300,000, 150,000 and 150,000 individuals from the three datasets respectively.

**Figure 1.**
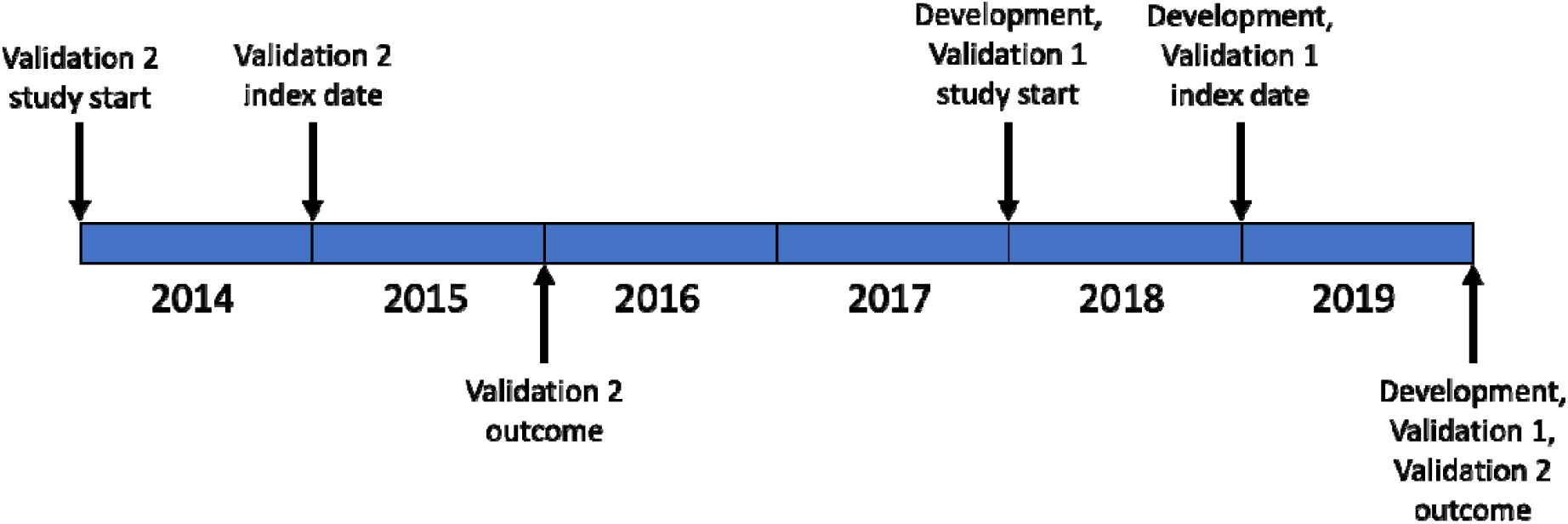
Study design.

We carefully curated the starting variables underlying the conditions used in the original development and validation, which was based on prior work on the epidemiology of multimorbidity in the UK [1, 11], with the same definitions and/or prescribing before the index date applied to SNOMED CT rather than to Read v2 (**S1 Table**). The exact same set of 46 starting variables was built using 66 variables within our Themes, Access, Dynamic Data Services (TADDS) library, and we retained the names from the original Cambridge Multimorbidity Score. We then applied the same logic and combined the anxiety and depression variables as described in Payne et al. [8] to yield 37 variables. Age and sex were included as covariates, with age censored at 95 years. For this study, we focused only on mortality as the outcome measure.

We extracted the following variables: pseudonymised practice and patient identifier (ID), sex, date of birth, date of death, dates of registration and deregistration with a general practice, and the 37 conditions.

### Statistical analyses

We constructed two time-to-mortality models using Cox proportional hazards in the development dataset. First, we performed a model with all 37 conditions as binary indicators, with sex, age (in 10 years) and a quadratic age term included as covariates. Then we ran a model using the 20 conditions that were considered the most important in Payne et al. [8] based on effect size, prevalence, and a combination of effect size and prevalence. Lastly, we conducted variable reduction by entering all predictors into a model, and then using backward elimination with the Akaike information criterion (AIC) [12] as the stopping criterion with the ‘fastbw’ function in the rms package.

We evaluated model discrimination using pseudo R-squared, Somers’ D, and Harrell’s C [13]. R-squared in a measure of explained variation in the model. Somers’ D quantifies the prognostic separation between observations with high and low predicted risk. Harrell’s C is the ratio of concordant pairs of observations to the number of comparable pairs; it estimates the concordance probability that larger predicted risks are associated with lower survival probabilities, when comparing the rankings of a pair of independent observations. Model calibration was assessed using a calibration curve, and recalibration was performed using resampling cross-validation to correct for overfitting with the ‘calibrate’ function in the rms package. Using model results from the development dataset, we then evaluated performance of the models in two validation datasets with synchronous and asynchronous outcomes (i.e. 1-year follow-up in the 2019 dataset, 1-year and 5-year follow-ups in the 2015 dataset).

All data preparation and analyses were conducted in R version 4.1.0 [14], using the following packages: ggplot2 (version 0.9.1) [15], lme4 (version 1.1-27) [16], lubridate (version 1.7.10) [17], randomizr (version 0.20.0) [18], rms (version 6.2-0) [19], survival (version 3.2-11) [20, 21], tableone (version 0.12.0) [22], and tidyverse (version 1.3.1) [23].

### Ethical considerations

A single comorbidity measure was required for our surveillance activity for Public Health England (PHE), its surveillance activities are now subsumed into the new UK Health Security Agency (UKHSA). Pseudonymised data for surveillance are extracted from volunteer general practices under Regulation 3 of the Health Service (Control of Patient Information) Regulations 2002 for health protection.

All potentially identifiable data were pseudonymised as close to source as possible and not made available to researchers; data were not extracted for patients who opted out of data sharing. All data are stored and processed at the Oxford-Royal College of General Practitioners Clinical Informatics Digital Hub (ORCHID), University of Oxford. This is listed by Health Data Research UK (HDRUK) as a trusted research environment and meets the standards of NHS Digital’s Data Security and Protection (DSP) toolkit (Organisation code: EE133863-MSD-NDPCHS).

## Results

The three datasets were generally comparable in distribution of age, sex, number of conditions and follow-up time (**Table 1**). Individuals in Validation Set 2 were slightly younger and healthier due to the earlier study index date.

**Table 1.** Descriptive statistics of the three datasets sampled from the Oxford-RCGP RSC cohort.

|  | Development<br>(2019) | Validation 1<br>(2019) | Validation 2<br>(2015) |
| --- | --- | --- | --- |
| Male | 148,672<br>(49.56%) | 74,463<br>(49.64%) | 74,527<br>(49.68%) |
| Age at index date, year |  |  |  |
| Mean ± sd | 48.44 ± 19.22 | 48.56 ± 19.39 | 47.84 ± 19.14 |
| Range | 16-95 | 16-95 | 16-95 |
| 65-84 years | 59,897<br>(19.97%) | 30,372<br>(20.25%) | 28,668<br>(19.11%) |
| ≥ 85 years | 9,390 (3.13%) | 4,844 (3.23%) | 4,558 (3.04%) |
| No. of conditions |  |  |  |
| Mean ± sd | 1.35 ± 1.85 | 1.37 ± 1.87 | 1.28 ± 1.78 |
| Range | 0-15 | 0-15 | 0-14 |
| 0 conditions | 138,076<br>(46.03%) | 68,928<br>(45.95%) | 71,635<br>(47.76%) |
| 1 condition | 66,053<br>(22.02%) | 32,377<br>(21.58%) | 32,400<br>(21.60%) |
| ≥ 2 conditions | 95,871<br>(31.96%) | 48,695<br>(32.46%) | 45,965<br>(30.64%) |
| No. of deaths in follow-up | 3,019 | 1,433 | 1,370 / 6,973 |
| Mean follow-up time (days) | 351.5 | 352.3 | 353.2 / 1564 |
| No. of people with complete follow-up | 278,494<br>(92.83%) | 139,670<br>(93.11%) | 140,513<br>(93.68%) /<br>109,612<br>(73.07%) |
| Total person-years* | 288,722.4 | 144,679.6 | 145,041.6 /<br>642,341.4 |
| Mortality rate (per 1,000 person-years) | 10.46 | 9.90 | 9.45 / 10.86 |

\* Calculated person-days then divided by 365.25

The prevalence of the included 37 conditions in the development dataset is presented in **Table 2**. Both the rates and the rankings show similar patterns to those observed in CPRD [8]. The top 20 conditions by prevalence and by effect size are listed in **S2 Table**.

**Table 2.** Prevalence of the 37 conditions in the development dataset, and the weights for the conditions included in the final model.

| Condition | Prevalence | Weight |
| --- | --- | --- |
| Hypertension | 62,854 (20.95%) |  |
| Anxiety or depression | 41,744 (13.91%) | 0.3242 |
| Painful condition | 41,461 (13.82%) | 0.4455 |
| Hearing loss | 27,083 (9.03%) |  |
| Asthma | 22,348 (7.45%) |  |
| Irritable bowel syndrome | 20,671 (6.89%) | -0.2037 |
| Diabetes | 20,232 (6.74%) | 0.2947 |
| Thyroid disorders | 18,732 (6.24%) |  |
| Coronary heart disease | 15,887 (5.30%) |  |
| Chronic kidney disease | 13,226 (4.41%) | 0.2137 |
| Diverticular disease of intestine | 10,502 (3.50%) |  |
| Disorder of prostate | 10,397 (3.47%) | -0.1878 |
| Atrial fibrillation | 9,105 (3.04%) | 0.3349 |
| Alcohol problems | 9,064 (3.02%) | 0.7922 |
| COPD | 7,542 (2.51%) | 0.7022 |
| Stroke & TIA | 7,415 (2.47%) |  |
| Rheumatoid arthritis | 7,352 (2.45%) |  |
| Constipation | 6,311 (2.10%) | 0.3830 |
| Cancer | 5,924 (1.97%) | 1.2026 |
| Peptic ulcer disease | 5,071 (1.69%) |  |
| Chronic sinusitis | 4,995 (1.67%) |  |
| Heart failure | 4,686 (1.56%) | 0.5052 |
| Psychoactive substance misuse | 4,139 (1.38%) | 0.4493 |
| Blindness & low vision | 3,823 (1.27%) |  |
| Dementia | 3,709 (1.24%) | 0.9380 |
| Psoriasis or eczema | 2,794 (0.93%) |  |
| Epilepsy | 2,580 (0.86%) | 0.4775 |
| Schizophrenia or bipolar disorder | 2,402 (0.80%) | 0.4825 |
| Inflammatory bowel disease | 2,371 (0.79%) |  |
| Chronic liver disease & viral hepatitis | 2,345 (0.78%) | 0.6862 |
| Anorexia or bulimia | 2,222 (0.74%) |  |
| Migraine | 1,594 (0.53%) |  |
| Bronchiectasis | 1,530 (0.51%) |  |
| Learning disability | 1,290 (0.43%) | 0.6373 |
| Parkinsonism | 920 (0.31%) | 0.5462 |
| Multiple sclerosis | 853 (0.28%) | 0.7616 |
| Peripheral vascular disease | 650 (0.22%) | 0.3346 |

Discrimination of 1-year mortality using the 37-condition model were high in both validation sets 1 and 2 (Harrell’s C = 0.92 for both models), and discrimination of 5-year mortality was only marginally worse in the validation dataset (c-index = 0.91) (**Table 3**). Prediction of 1-year and 5-year mortality using the original simplified 20-condition model showed a similar pattern.

**Table 3.** Model discrimination, as assessed using pseudo R-squared, Somers’ D and Harrell’s C.

|  | 37-condition | 20-condition | Reduced model |
| --- | --- | --- | --- |
| *Pseudo R-squared* | 0.153 | 0.152 | 0.153 |
| *Somers' D* | 0.851 | 0.847 | 0.851 |
| *Harrell's C* |  |  |  |
| Development | 0.9253<br>(se = 0.0022) | 0.9236<br>(se = 0.0022) | 0.9255<br>(se = 0.0021) |
| Validation 1 | 0.9200<br>(se = 0.0035) | 0.9184<br>(se = 0.0035) | 0.9206<br>(se = 0.0035) |
| Validation 2, 1-year follow-up | 0.9204<br>(se = 0.0033) | 0.9182<br>(se = 0.0033) | 0.9203<br>(se = 0.0033) |
| Validation 2, 5-year follow-up | 0.9071<br>(se = 0.0016) | 0.9055<br>(se = 0.0016) | 0.9072<br>(se = 0.0016) |

Our reduced model retained 21 conditions, which partly overlapped with those in the 20-condition model (**Table 4**), and showed similar performance. The model had reasonable calibration, although it was found to under-predict survival at lower risks (<60%). Much of this under prediction was removed in predictions adjusted for overfitting (**Figure 2**).

**Table 4.**
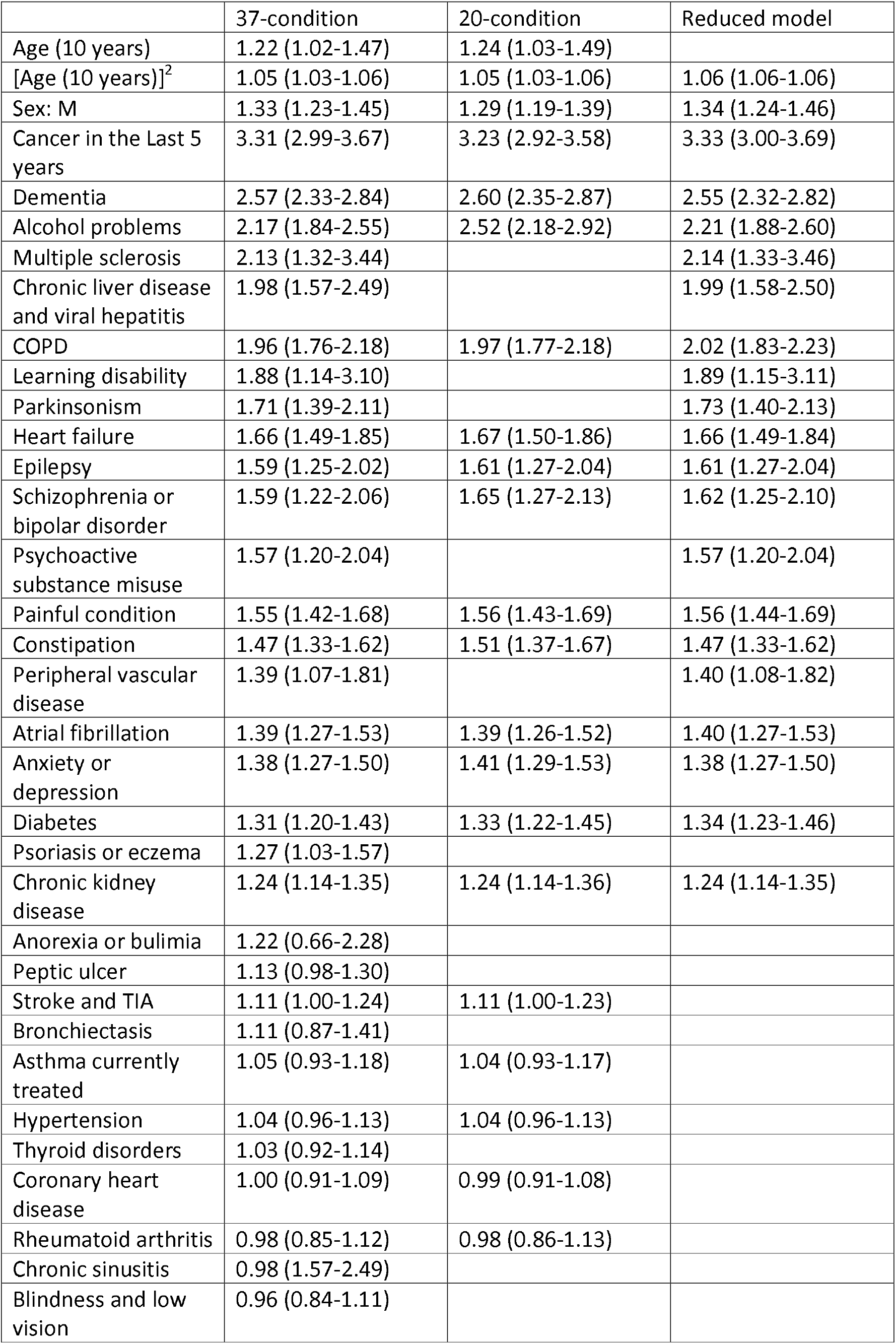
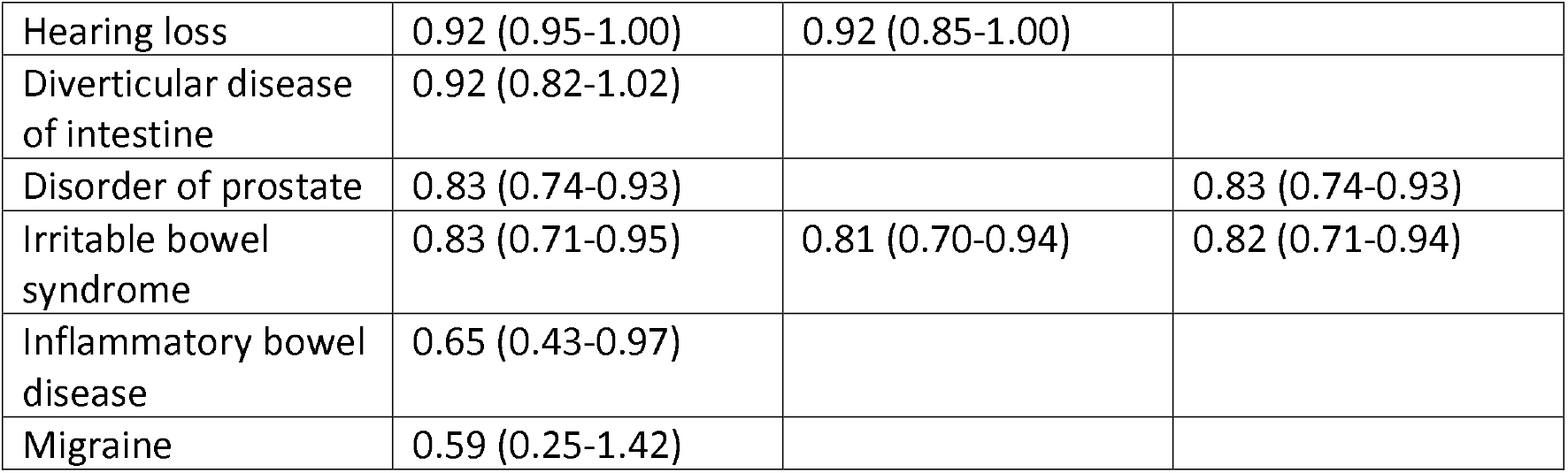
Hazard ratios (95% CIs) of the predictors from the three models.

|  | 37-condition | 20-condition | Reduced model |
| --- | --- | --- | --- |
| Age (10 years) | 1.22 (1.02-1.47) | 1.24 (1.03-1.49) |  |
| [Age (10 years)]<sup>2</sup> | 1.05 (1.03-1.06) | 1.05 (1.03-1.06) | 1.06 (1.06-1.06) |
| Sex: M | 1.33 (1.23-1.45) | 1.29 (1.19-1.39) | 1.34 (1.24-1.46) |
| Cancer in the Last 5 years | 3.31 (2.99-3.67) | 3.23 (2.92-3.58) | 3.33 (3.00-3.69) |
| Dementia | 2.57 (2.33-2.84) | 2.60 (2.35-2.87) | 2.55 (2.32-2.82) |
| Alcohol problems | 2.17 (1.84-2.55) | 2.52 (2.18-2.92) | 2.21 (1.88-2.60) |
| Multiple sclerosis | 2.13 (1.32-3.44) |  | 2.14 (1.33-3.46) |
| Chronic liver disease and viral hepatitis | 1.98 (1.57-2.49) |  | 1.99 (1.58-2.50) |
| COPD | 1.96 (1.76-2.18) | 1.97 (1.77-2.18) | 2.02 (1.83-2.23) |
| Learning disability | 1.88 (1.14-3.10) |  | 1.89 (1.15-3.11) |
| Parkinsonism | 1.71 (1.39-2.11) |  | 1.73 (1.40-2.13) |
| Heart failure | 1.66 (1.49-1.85) | 1.67 (1.50-1.86) | 1.66 (1.49-1.84) |
| Epilepsy | 1.59 (1.25-2.02) | 1.61 (1.27-2.04) | 1.61 (1.27-2.04) |
| Schizophrenia or bipolar disorder | 1.59 (1.22-2.06) | 1.65 (1.27-2.13) | 1.62 (1.25-2.10) |
| Psychoactive substance misuse | 1.57 (1.20-2.04) |  | 1.57 (1.20-2.04) |
| Painful condition | 1.55 (1.42-1.68) | 1.56 (1.43-1.69) | 1.56 (1.44-1.69) |
| Constipation | 1.47 (1.33-1.62) | 1.51 (1.37-1.67) | 1.47 (1.33-1.62) |
| Peripheral vascular disease | 1.39 (1.07-1.81) |  | 1.40 (1.08-1.82) |
| Atrial fibrillation | 1.39 (1.27-1.53) | 1.39 (1.26-1.52) | 1.40 (1.27-1.53) |
| Anxiety or depression | 1.38 (1.27-1.50) | 1.41 (1.29-1.53) | 1.38 (1.27-1.50) |
| Diabetes | 1.31 (1.20-1.43) | 1.33 (1.22-1.45) | 1.34 (1.23-1.46) |
| Psoriasis or eczema | 1.27 (1.03-1.57) |  |  |
| Chronic kidney disease | 1.24 (1.14-1.35) | 1.24 (1.14-1.36) | 1.24 (1.14-1.35) |
| Anorexia or bulimia | 1.22 (0.66-2.28) |  |  |
| Peptic ulcer | 1.13 (0.98-1.30) |  |  |
| Stroke and TIA | 1.11 (1.00-1.24) | 1.11 (1.00-1.23) |  |
| Bronchiectasis | 1.11 (0.87-1.41) |  |  |
| Asthma currently treated | 1.05 (0.93-1.18) | 1.04 (0.93-1.17) |  |
| Hypertension | 1.04 (0.96-1.13) | 1.04 (0.96-1.13) |  |
| Thyroid disorders | 1.03 (0.92-1.14) |  |  |
| Coronary heart disease | 1.00 (0.91-1.09) | 0.99 (0.91-1.08) |  |
| Rheumatoid arthritis | 0.98 (0.85-1.12) | 0.98 (0.86-1.13) |  |
| Chronic sinusitis | 0.98 (1.57-2.49) |  |  |
| Blindness and low vision | 0.96 (0.84-1.11) |  |  |
| Hearing loss | 0.92 (0.95-1.00) | 0.92 (0.85-1.00) |  |
| Diverticular disease of intestine | 0.92 (0.82-1.02) |  |  |
| Disorder of prostate | 0.83 (0.74-0.93) |  | 0.83 (0.74-0.93) |
| Irritable bowel syndrome | 0.83 (0.71-0.95) | 0.81 (0.70-0.94) | 0.82 (0.71-0.94) |
| Inflammatory bowel disease | 0.65 (0.43-0.97) |  |  |
| Migraine | 0.59 (0.25-1.42) |  |  |

**Figure 2.**
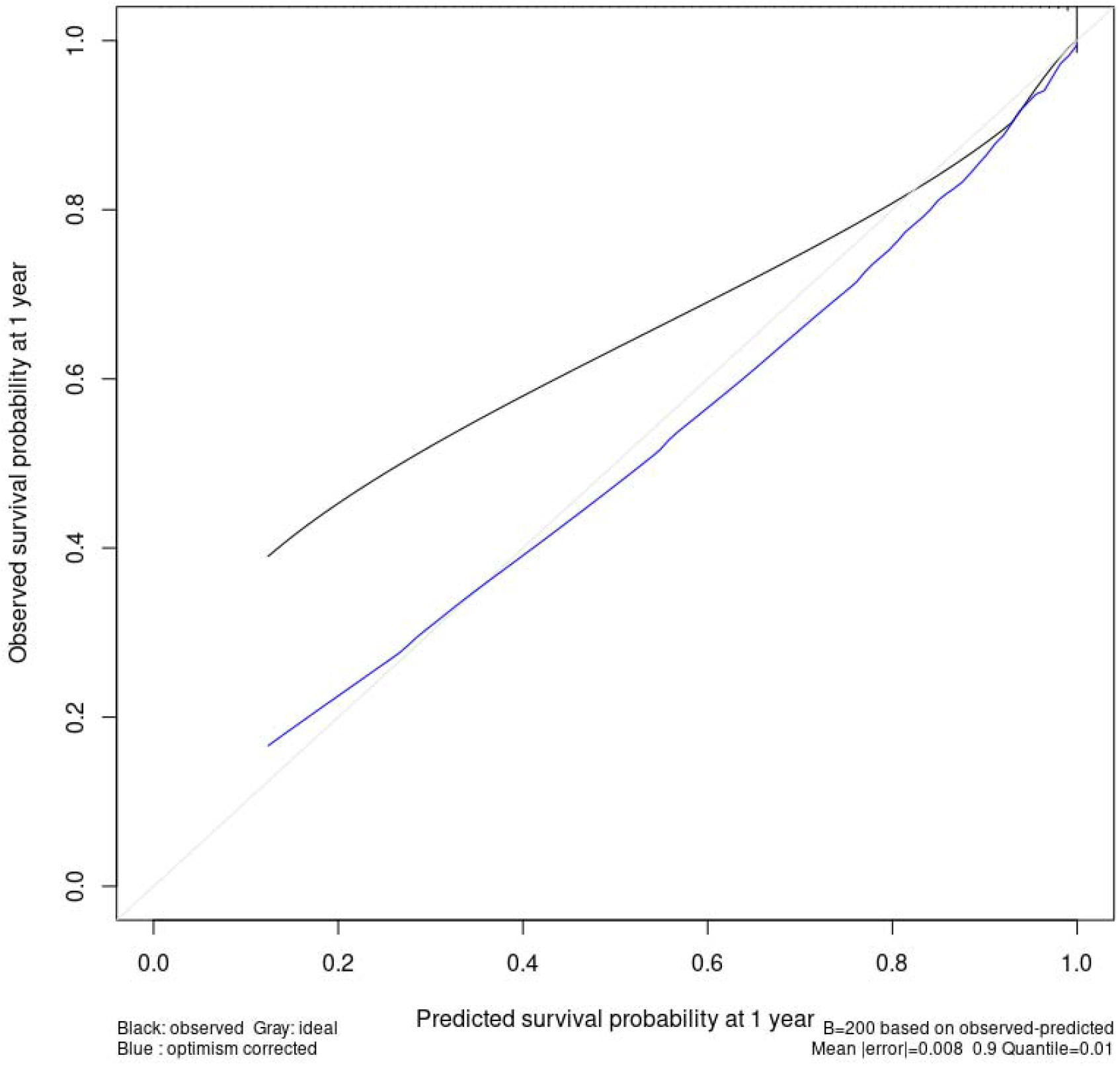
Calibration curve for the 21-condition model.

## Discussion

### Key findings

In this study, we developed and validated a modified version of the CMMS to be used in a wider age range and using SNOMED CT. Our reduced 21-condition model performed similarly to both the full model and the original 20-condition model in predicting mortality with excellent discrimination and reasonable calibration. We have opted to use the unadjusted 21-condition model as this would maximise its use in studies of different designs, where researchers can apply their own adjustments for age and sex. We plan to use this multimorbidity score in our epidemiological studies (including COVID-19 studies), and to make this available to the wider international SNOMED CT community.

### Comparison with the literature

A number of comorbidity indices and adaptations have been developed in administrative data studies, which are either solely diagnosis-based or solely medication-based [24]. Our score uses a different approach that combines information from clinical terms as well as prescriptions, and additionally includes a 12-month timeframe in the definition of certain conditions. This allows the severity and/or recency of some conditions (e.g. constipation, cancer) to be taken into account in the calculation of the score.

Our study retained a slightly different set of conditions in our reduced model to Payne et al. [8]. Since variable reduction in our model was based on AIC rather than the combination of effect size and prevalence, our modified score included some less prevalent conditions that are strongly associated with mortality such as multiple sclerosis and learning disability. The differences in included conditions and weightings between our model and Payne et al.’s [8] may also be partly explained by age group differences in multimorbidity patterns as we used a lower age cut-off of 16 years. Earlier research has shown while multimorbidity in later life tends to involve multiple ‘concordant’ conditions (typically vascular and metabolic conditions), multimorbidity in earlier adulthood generally involves a mix of physical and mental conditions [25].

### Strengths and limitations

Our study used a large, up-to-date, nationally representative cohort, which included all patients aged 16 years and over, and our results were validated using both synchronous and asynchronous datasets. Our analysis was based on SNOMED CT, now used across English General Practice as well as internationally. We believe the results are generalisable to other cohorts and potentially other countries that use similarly coded primary care data.

We derived CMMS weights only for mortality but not unplanned hospital admissions or primary care consultations. Mortality tends to be the most commonly used outcome in the development of comorbidity indices in the literature [24], and we felt that having only one set of weights would allow it to be easier to apply and to interpret in different datasets.

The list of conditions used in this study were exactly as included in the original development and validation of the CMMS by Payne et al. [8], which was based on earlier literature on multimorbidity in primary care [1, 11]. These studies did not include other common conditions that might be expected to be included in other multimorbidity indices or that are highly clinically relevant (e.g. obesity).

## Conclusion

In this study we described the development and validation of a modified version of the CMMS for predicting mortality. The inclusion of a wider age range may improve the generalisability of the score over the original. Because it is based on SNOMED CT rather than Read codes it is applicable to today’s English General Practice data and should also increase its applicability in other contexts.

## Data Availability

The Oxford-RCGP RSC dataset can be accessed by researchers; approval is on a project-by-project basis (orchid.phc.ox.ac.uk/index.php/rcgp-rsc/). All patient-level data used was pseudonymised and in compliance with information governance policies. The analysis of the data was carried out in the secure servers at the University of Oxford. Patient-level data cannot be taken out of the secure network.

## Acknowledgements

JPS is funded by the Wellcome Trust/Royal Society via a Sir Henry Dale Fellowship (ref: 211182/Z/18/Z). JPS also receives funding via an NIHR Oxford Biomedical Research Centre (BRC) Senior Fellowship. For the purpose of Open Access, the author has applied a CC BY public copyright licence to any Author Accepted Manuscript version arising from this submission. This publication presents independent research supported by the National Institute for Health Research (NIHR). The views expressed are those of the author(s) and not necessarily those of the NHS, the NIHR or the Department of Health and Social Care.

## Data availability

The Oxford-RCGP RSC dataset can be accessed by researchers; approval is on a project-by-project basis (orchid.phc.ox.ac.uk/index.php/rcgp-rsc/). Ethical approval by an NHS Research Ethics Committee/other appropriate approval is needed before any data release. Researchers wishing to directly analyse patient-level pseudonymised data will be required to complete information governance training and work on the data from the secure servers at the University of Oxford. Patient-level data cannot be taken out of the secure network.

---

**S1 Figure.**
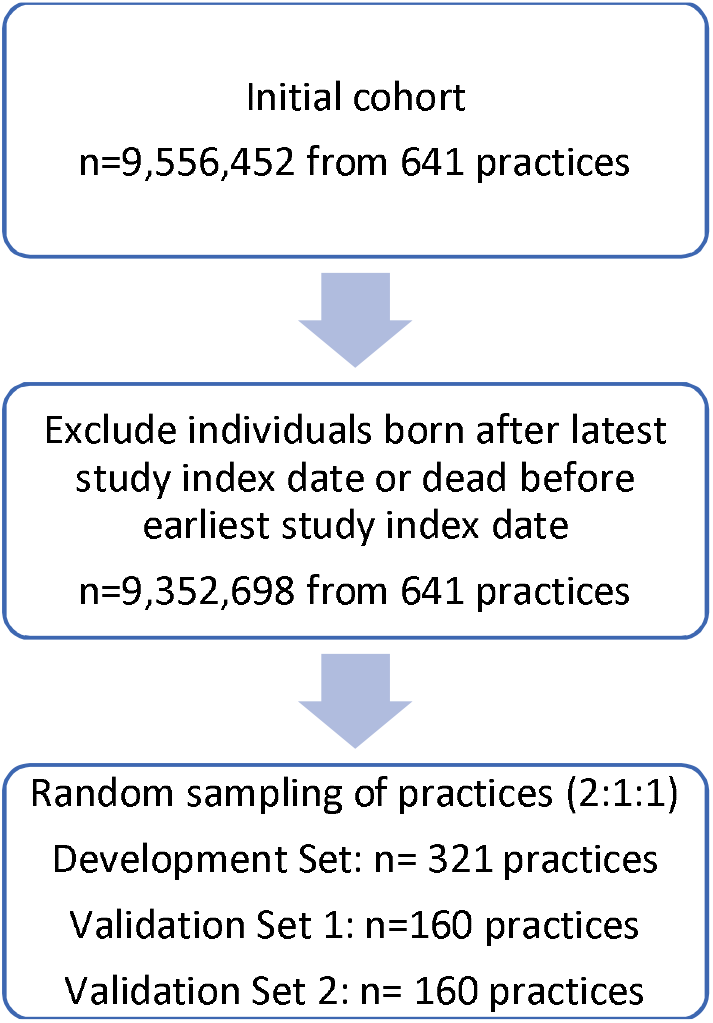
Flow diagram for selection of practices. A measure of mortality not explained by age (standardised) or sex was computed for each practice (i.e. best linear unbiased predictor from a mixed effects logistic regression with a practice random effect), which was sorted into four bins using mean – sd, mean, mean + sd and then block randomised into three datasets.

**S2 Figure.**
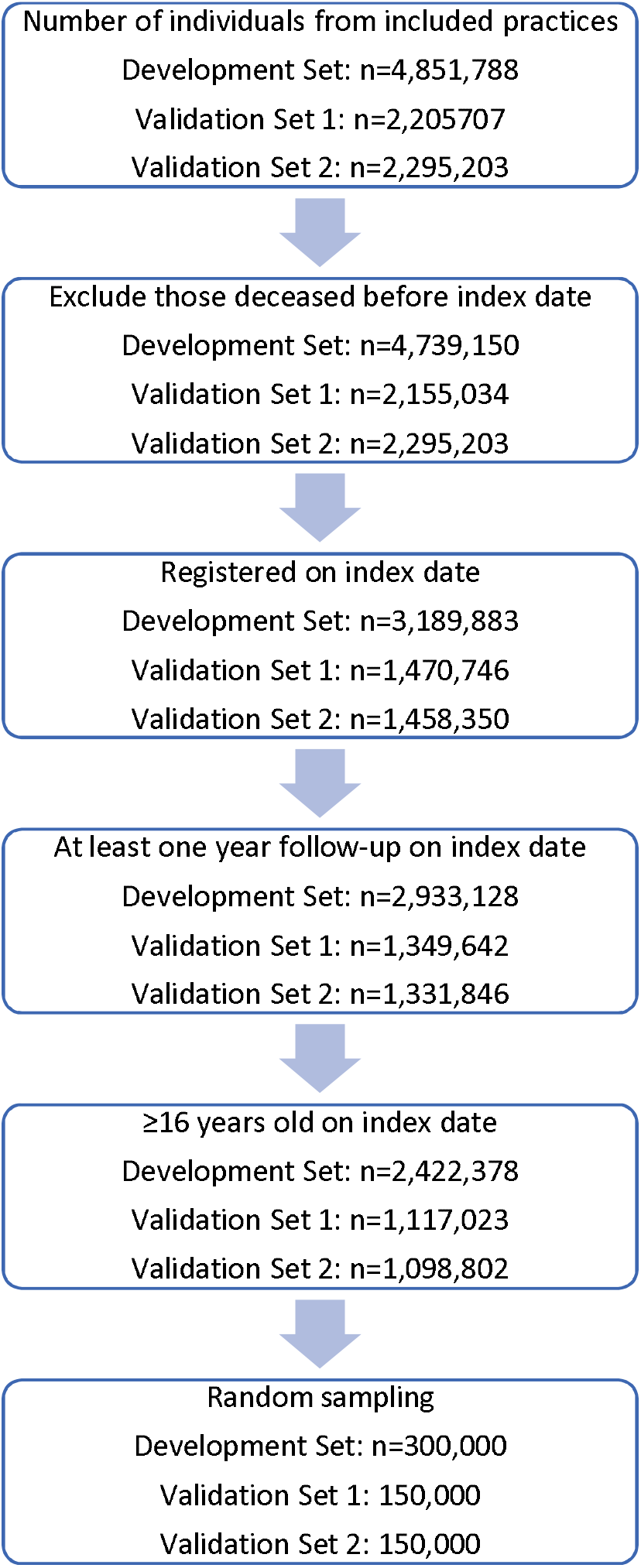
Flow diagram for selection of individuals.

**S1 Table.** List of the 37 morbidities included in the baseline model (adapted from Payne et al. (2020)).

|  |  |
| --- | --- |
| Morbidities based on SNOMED CT ever recorded |  |
| Alcohol problems | Heart failure |
| Anorexia or bulimia | Hypertension |
| Atrial fibrillation | Inflammatory bowel disease |
| Blindness and low vision | Learning disability |
| Bronchiectasis | Multiple sclerosis |
| Chronic liver disease and viral hepatitis | Parkinsonism |
| Chronic sinusitis | Peptic ulcer disease |
| COPD | Peripheral vascular disease |
| Coronary heart disease | Disorder of prostate |
| Dementia | Psychoactive substance misuse (not alcohol) |
| Diabetes | Rheumatoid arthritis |
| Diverticular disease of intestine | Stroke & transient ischaemic attack |
| Hearing loss | Thyroid disorders |
| Morbidities based on prescription in last 12 months |  |
| Constipation (currently treated) | ≥4 laxative prescriptions |
| Migraine | ≥4 prescription-only medicine anti-migraine prescriptions |
| Morbidities based on combination of SNOMED CT ever recorded and/or prescription in last 12 months |  |
| Epilepsy (currently treated) | SNOMED CT AND ≥1 antiepileptic prescription |
| Asthma (currently treated) | SNOMED CT AND ≥1 asthma prescription |
| Irritable bowel syndrome | SNOMED CT OR ≥4 antispasmodic prescriptions |
| Psoriasis or eczema | SNOMED CT AND ≥4 related prescriptions (excluding simple emollients) |
| Other morbidities |  |
| Anxiety or depression | SNOMED CT (anxiety or depression) in last 12 months OR ≥4 anxiolytic/hypnotic prescriptions in last 12 months OR ≥4 anti-depressant prescriptions (excluding low dose tricyclics) in last 12 months |
| (New) cancer in last 5 years (excluding non-melanoma skin cancer) | SNOMED CT (first) recorded in last 5 years |
| Chronic kidney disease | Highest value of last 2 eGFR readings is <60 ml/min |
| Painful condition | ≥4 prescription-only medicine analgesics in last 12 months OR (≥4 specified anti-epileptics in last 12 months AND no epilepsy SNOMED CT ever recorded) |
| Schizophrenia or bipolar disorder | SNOMED CT ever recorded OR lithium ever prescribed |

**S2 Table.** Top 20 conditions by prevalence and by effect size in the development dataset.

| By prevalence | By absolute effect size |
| --- | --- |
| Hypertension | Dementia |
| Anxiety or depression | Cancer in the last 5 years |
| Painful condition | Migraine |
| Hearing loss | Painful condition |
| Asthma | Chronic kidney disease |
| Irritable bowel syndrome | COPD |
| Diabetes | Parkinsonism |
| Thyroid disorders | Constipation |
| Coronary heart disease | Atrial fibrillation |
| Chronic kidney disease | Hypertension |
| Diverticular disease of intestine | Heart failure |
| Disorder of prostate | Chronic liver disease and viral hepatitis |
| Atrial fibrillation | Alcohol problems |
| Alcohol problems | Inflammatory bowel disease |
| COPD | Epilepsy |
| Stroke and transient ischaemic attack | Irritable bowel syndrome |
| Rheumatoid arthritis | Peripheral vascular disease |
| Constipation | Hearing loss |
| Cancer in the last 5 years | Schizophrenia or bipolar disorder |
| Peptic ulcer disease | Bronchiectasis |

## Notes

### Competing Interest Statement

The authors have declared no competing interest.

### Author Declarations

Ethics committee of University of Oxford waived ethical approval for this work

## References

1. Cassell A, Edwards D, Harshfield A, Rhodes K, Brimicombe J, Payne R, et al. The epidemiology of multimorbidity in primary care: a retrospective cohort study. British Journal of General Practice. 2018;68(669):e245. doi: https://doi.org/10.3399/bjgp18X695465.

2. Charlson ME, Pompei P, Ales KL, MacKenzie CR. A new method of classifying prognostic comorbidity in longitudinal studies: Development and validation. Journal of Chronic Diseases. 1987;40(5):373–83. doi: https://doi.org/10.1016/0021-9681(87)90171-8.

3. Elixhauser A, Steiner C, Harris DR, Coffey RM. Comorbidity Measures for Use with Administrative Data. Medical Care. 1998;36(1). doi: https://doi.org/10.1097/00005650-199801000-00004.

4. Deyo RA, Cherkin DC, Ciol MA. Adapting a clinical comorbidity index for use with ICD-9-CM administrative databases. Journal of Clinical Epidemiology. 1992;45(6):613–9. doi: https://doi.org/10.1016/0895-4356(92)90133-8.

5. Sundararajan V, Henderson T, Perry C, Muggivan A, Quan H, Ghali WA. New ICD-10 version of the Charlson comorbidity index predicted in-hospital mortality. Journal of Clinical Epidemiology. 2004;57(12):1288–94. doi: https://doi.org/10.1016/j.jclinepi.2004.03.012.

6. Palella FJ, J., Delaney KM, Moorman AC, Loveless MO, Fuhrer J, Satten GA, et al. Declining Morbidity and Mortality among Patients with Advanced Human Immunodeficiency Virus Infection. New England Journal of Medicine. 1998;338(13):853–60. doi: https://doi.org/10.1056/NEJM199803263381301.

7. Zavascki AP, Fuchs SC. The need for reappraisal of AIDS score weight of Charlson comorbidity index. Journal of Clinical Epidemiology. 2007;60(9):867–8. doi: https://doi.org/10.1016/j.jclinepi.2006.11.004.

8. Payne RA, Mendonca SC, Elliott MN, Saunders CL, Edwards DA, Marshall M, et al. Development and validation of the Cambridge Multimorbidity Score. Canadian Medical Association Journal. 2020;192(5):E107. doi: https://doi.org/10.1503/cmaj.190757.

9. NHS Digital. Read Codes 2020 [26/07/2021]. Available from: https://digital.nhs.uk/services/terminology-and-classifications/read-codes.

10. NHS Digital. SNOMED CT 2021 [26/07/2021]. Available from: https://digital.nhs.uk/services/terminology-and-classifications/snomed-ct.

11. Barnett K, Mercer SW, Norbury M, Watt G, Wyke S, Guthrie B. Epidemiology of multimorbidity and implications for health care, research, and medical education: a cross-sectional study. The Lancet. 2012;380(9836):37–43. doi: 10.1016/S0140-6736(12)60240-2.

12. Akaike H. A new look at the statistical model identification. IEEE Transactions on Automatic Control. 1974;19(6):716–23. doi: https://doi.org/10.1109/TAC.1974.1100705.

13. Harrell FE, Jr., Califf RM, Pryor DB, Lee KL, Rosati RA. Evaluating the Yield of Medical Tests. JAMA. 1982;247(18):2543–6. doi: https://doi.org/10.1001/jama.1982.03320430047030.

14. R Core Team. R: A language and environment for statistical computing. Vienna, Austria: R Foundation for Statistical Computing; 2021.

15. Wickham H. ggplot2: Elegant Graphics for Data Analysis. New York: Springer-Verlag; 2016.

16. Bates D, Mächler M, Bolker B, Walker S. Fitting linear mixed-effects models using lme4. Journal of Statistical Software. 2015;67(1):1–48. doi: https://doi.org/10.18637/jss.v067.i01.

17. Grolemund G, Wickham H. Dates and times made easy with lubridate. Journal of Statistical Software. 2011;40(3):1–25.

18. Coppock A. randomizr: Easy-to-Use Tools for Common Forms of Random Assignment and Sampling. R package version 0.20.0. 2019. Available from: https://CRAN.R-project.org/package=randomizr.

19. Harrell FE, Jr. rms: Regression Modeling Strategies. R package version 6.2-0 2021. Available from: https://CRAN.R-project.org/package=rms.

20. Therneau T. A Package for Survival Analysis in R. R package version 3.2-11. 2021. Available from: https://CRAN.R-project.org/package=survival.

21. Therneau TM, Grambsch PM. Modeling Survival Data: Extending the Cox Model. New York: Springer; 2000.

22. Yoshida K, Bartel A. tableone: Create ‘Table 1’ to Describe Baseline Characteristics with or without Propensity Score Weights. R package version 0.12.0. 2020. Available from: https://CRAN.R-project.org/package=tableone.

23. Wickham H, Averick M, Bryan J, Chang W, D’Agostino McGowan L, François R, et al. Welcome to the tidyverse. Journal of Open Source Software. 2019;4(43):1686. doi: https://doi.org/10.21105/joss.01686.

24. Yurkovich M, Avina-Zubieta JA, Thomas J, Gorenchtein M, Lacaille D. A systematic review identifies valid comorbidity indices derived from administrative health data. Journal of Clinical Epidemiology. 2015;68(1):3–14. doi: 10.1016/j.jclinepi.2014.09.010.

25. McLean G, Gunn J, Wyke S, Guthrie B, Watt GCM, Blane DN, et al. The influence of socioeconomic deprivation on multimorbidity at different ages: a cross-sectional study. British Journal of General Practice. 2014;64(624):e440. doi: 10.3399/bjgp14X680545.

